# Deep generative models for vessel segmentation in CT angiography of the brain

**DOI:** 10.1101/2025.03.07.25322919

**Authors:** Henk van Voorst, Jiahang Su, Praneeta R. Konduri, Charles B.L.M. Majoie, Yvo B.W.E.M. Roos, Bart J. Emmer, Henk A. Marquering, Bob D. de Vos, Matthan W.A. Caan, Ivana Išgum MR CLEAN Registry collaborators

**Affiliations:** Department of Radiology and Nuclear Medicine, Amsterdam UMC, Amsterdam, The Netherlands; Department of Biomedical Engineering and Physics, Amsterdam UMC, Amsterdam, The Netherlands; Department of Radiology, Erasmus MC, Rotterdam, The Netherlands; Department of Neurology, Amsterdam UMC, Amsterdam, The Netherlands; Informatics Institute, Faculty of Science, University of Amsterdam, Amsterdam, The Netherlands

**Keywords:** Segmentation, Vessels, Generative adversarial network, CT angiography, Ischemic stroke

## Abstract

Automated vessel segmentation in brain CT angiography (CTA) remains challenging despite the potential benefit of applications. Expert acquisition of reference vessel segmentations is a laborious task. We propose an unsupervised generative deep learning approach that can be trained for vessel segmentation in brain CTA using a large dataset (n=908) of unlabelled brain CTAs and non-contrast enhanced CTs (NCCTs). Our unsupervised approach uses a conditional generative adversarial network (GAN) for CTA to NCCT translation by generating a contrast map that allows for automatic extraction of vessel segmentations. Furthermore, we propose a 3D Frangi filter-based loss function to enhance tubular structures in the contrast map to improve vessel segmentations. We used a hold-out test set of 9 CTA volumes with manually annotated reference segmentations. We compared our unsupervised approach with a state-of-the-art supervised nnUnet, trained and evaluated with test set using 9-fold nested cross-validation. Evaluation metrics included voxel-wise Dice similarity coefficient (DSC), true positive rate (TPR), and false positive rate (FPR). The DSC was 4% lower for the unsupervised approach (DSC: 0.74) compared to the supervised nnUnet (DSC: 0.78). Both the TPR and FPR were higher for the unsupervised approach (TPR: 0.75, FPR/1000 voxels:2.05) compared to the supervised nnUnet (TPR:0.71, FPR/1000 voxels:0.87). Hence, the quantitative results showed that our unsupervised method approaches a supervised state-of-the-art segmentation network. The results demonstrate that an unsupervised generative deep learning approach for the segmentation of intracranial vessels is feasible without laborious manual segmentations.

**Highlights:**

- To train supervised segmentation models laborious manual segmentations are needed
- Unsupervised generative deep learning does not require manual segmentations
- Our unsupervised method combines L1, adversarial, and a novel Frangiloss
- Varying loss function combinations can reduce false positives or false negatives
- Our method approached the performance of a state-of-the-art supervised nnUnet

## 1. Introduction

Segmentation of the vessel lumen in CT angiography (CTA) of the brain can be used to support radiologists in the diagnosis, prognosis, and treatment of cerebrovascular diseases [1, 2, 3]. Specifically, in patients with ischemic stroke, automated segmentation of vessels in the brain can be used to expedite the detection and localization of occlusions, assess the arterial collateral circulation to the infarcted area, and assess the venous outflow from the infarcted area [1, 2, 3]. Therefore, numerous methods for automated vessel segmentation in CTA and MRA have been developed [4, 5].

Brain vessels appear as thin tubular structures in an image and present similar segmentation problems in MRA and CTA. Moreover, their intensity is increased in CTA due to intravenously administered angiographic contrast fluid and in MRA due to the magnetic properties of the flowing blood or injected contrast fluid. The intensity and shape characteristics of brain vessels were first exploited by conventional rule-based programming segmentation techniques such as tracking algorithms, surface detection algorithms, and algorithms using pre-defined convolution filters [4, 5]. Manniesing et al. used a tracking algorithm based on surface evolution to exploit the connectivity of the vessel’s voxels in an image to construct or refine a vessel centerline from the manually or automatically defined seed points, which is subsequently used to identify vessel boundary using a surface detection algorithm [6]. Centerline tracking and surface detection algorithms result in accurate vessel segmentation for vessels with large diameters and when vessel trees have a clear start and end point [6, 7]. However, the segmentation performance of these methods is challenged by branching vessels and circular vessel networks that occur in the brain, such as the Circle of Willis (CoW). As a result, most tracking algorithms for vessel segmentation in the brain would require a large burden for the user due to manual tasks such as manual seed-point selection [7]. Contrarily, vessel segmentation techniques relying on predefined filtering techniques can be applied to a whole image without manual interaction. For example, Hassanpour et al. used pre-defined convolution filters to perform morphological operations for vessel segmentation [8]. However, the pre-defined convolution filters must be fine-tuned and refined to achieve good performance using postprocessing steps specific to a dataset [8].

Alternatively, local features can be extracted and combined with an algorithm or rule-based method to classify the presence of a vessel. Frangi et al. developed the Frangi filter, which utilizes eigenvalue ratios of the local intensity changes measured with Hessian filtering to construct vesselness features [9, 10]. Three vesselness features are combined by attributing predefined relative weights to segment vessels. Lu et al. replaced the pre-defined weights with an expectation-maximization algorithm and probabilistic noise filtering to construct a vessel segmentation algorithm that can be adapted to variations between images. Although conventional methods result in high-quality vessel segmentation on brain MRA [4, 5], these approaches typically suffer from false positive segmentation of other high-intensity areas in CTA such as bone and (arterial) calcifications and therefore require further post-processing [10, 6, 4, 5]. Furthermore, conventional methods are tailored to a specific dataset and may suffer from limited generalizability. Regularly appearing variations in contrast attenuation and noise levels in a scan between patients and between scanners may degrade the performance of these conventional vessel segmentation methods [11].

The introduction of supervised deep learning approaches using convolution neural networks (CNNs) has led to remarkable performance gains for image segmentation and they are currently considered the state-of-the-art for segmentation of the vessels in the brain [4, 5]. Specifically, the Unet architecture that combines several down- and up-sampling convolutions with skip connections to enable the use of features with varying levels of abstraction and receptive fields has become the first-choice segmentation method for various applications [12]. The original 2D Unet architecture for vessel segmentation was extended by Hilbert et al. and Tetteh et al. to 3D approaches, improving segmentation quality in 3D imaging such as brain MRA and CTA [13, 14]. Whereas supervised deep learning models for vessel segmentation in MRA achieve accurate segmentation [13, 14], attaining highly accurate segmentations for CTA remains challenging [2, 14, 15, 16, 17, 18]. More recently, Liu et al. [19] and Su et al. [20] have considered vessel centerline extraction for brain CTA instead of segmenting the lumen. Although these centerline extraction techniques have supported lumen segmentation in the coronary arteries and aorta [21], to date, lumen segmentation leveraged by centerline extraction has not been studied in brain vessels. Irrespective of the use of centerlines extraction techniques to segment the vessel lumen, supervised deep learning techniques require manual annotations as reference for training, which are extremely time-consuming to acquire [22, 2]. Hence, alternative approaches that require fewer or no manually annotated vessels for training vessel segmentation models would allow for the use of larger unannotated training datasets and would reduce the annotation burden.

In contrast to supervised methods, unsupervised deep learning models do not require explicit reference annotations for training. Specifically, image-to-image translation methods that transform an input image from one domain (source domain) to an image from another domain (target domain) have gained significant interest after the introduction of generative adversarial networks (GANs) [23]. GANs contain at least two CNNs: a generator CNN to transform an image and a discriminator CNN that classifies if images are synthesized or real to provide feedback to the generator. Most segmentation methods using a GAN consider the GAN losses in addition to a supervised loss function, which would still require manual annotations [24]. For example, Chen et al. refined supervised deep learning-based vessel segmentation using a GAN by combining the input MRA with predicted or manual reference segmentations [25]. Huang et al. used a 3D Unet optimized jointly with a binary cross-entropy between predicted segmentations in CTA and self-generated reference segmentations in combination with an L1-loss for CTA to NCCT translation [18]. While Huang et al. demonstrated good performance on single-center data, generalization to multi-center settings is likely challenged by varying acquisition protocols, noise levels in scans, and contrast levels due to the timing of CTA acquisition [11]. To address the challenges of manual annotation of brain vessels, both arteries and veins, required for supervised deep learning, we propose an unsupervised generative deep learning approach for 3D vessel segmentation in CTA. We exploit the difference in contrast attenuation between CTA and NCCT and train a GAN that removes contrast from a CTA to generate a synthetic NCCT. For this, the generator CNN synthesizes a contrast map (Cmap), which is then subtracted from the CTA, resulting in a synthetic NCCT. The discriminator CNN classifies whether the NCCT is synthesized (NCCT_syn_) or real (NCCT_real_). Based on the discriminator classification, the adversarial loss (Adv) and the mean absolute difference between the (NCCT_syn_) and (NCCT_real_) (L1-loss), the (NCCT_syn_) is optimized. By optimizing the (NCCT_syn_) the obtained contrast map is refined to remove contrast in the CTA. In addition to the adversarial and L1 losses that focus on CTA to NCCT translation, a 3D Frangi-loss is introduced to enhance vessels and reduce noise in the contrast map during training. Finally, the contrast map is used to extract vessel segmentations. We train and evaluate the method with a nationwide multi-center dataset.

## 2. Data

We included data from the Multicenter collaboration for endovascular treatment of acute ischemic stroke in the Netherlands (MR CLEAN) Registry [26]. In the Registry, patients (n=3279) were included between March 2014 and November 2017 from 16 hospitals in the Netherlands. According to European guidelines for acute stroke [27], NCCT and CTA were acquired consecutively to rule out hemorrhagic stroke and to diagnose ischemic stroke by detecting an occlusion. We excluded patients if thin slice NCCT and CTA (slice thickness ≤ 1.0*mm*) were unavailable in the database or if image quality was poor per a visual check (n excluded=2213). All patients were imaged and included once in our data. Details of in and exclusion are presented in Figure 1.

**Figure 1:**
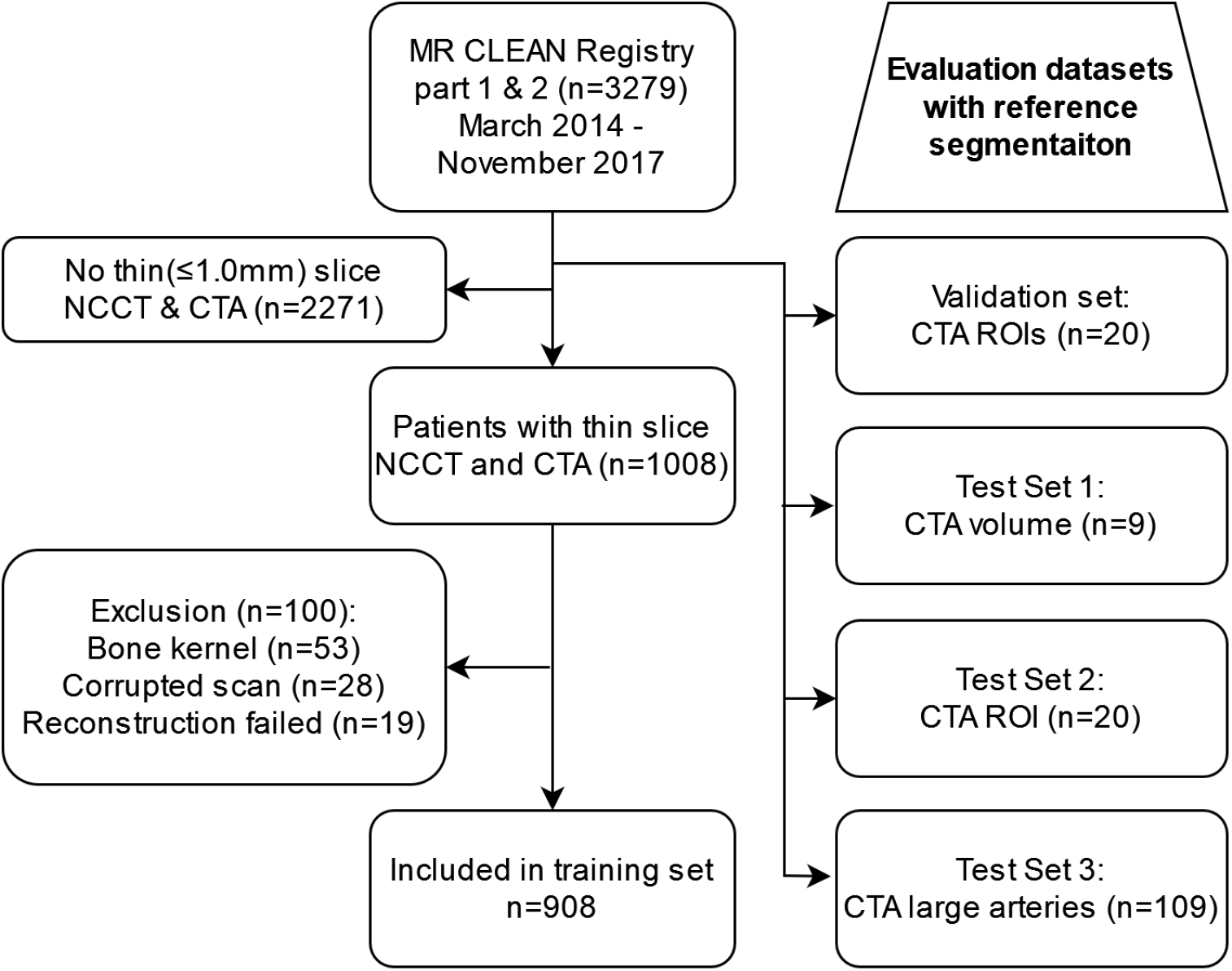
Patient inclusion

### 2.1. Training, validation, and test sets

In Figure 1 we describe the brain imaging from 1066 patients that we used for this study; a training set of consecutive NCCT and CTA per patient (n=908), a validation set with defined regions of interest from CTA (ROI-val) (n=20), a CTA volume test set (Set 1, n=9), a CTA ROI test set similar to the validation set (Set 2, n=20), and 109 CTAs of the large arteries (Set 3). Only the validation and test set CTAs have manual reference vessel segmentations for evaluation, training data does not have reference segmentations. Regions of interest (ROI) are sized 128×128×128 voxels. Expert radiologists manually selected the ROIs to represent a realistic variation in contrast phase and noise levels that occur in acute ischemic stroke patients and then randomly split between validation and test set [2]. Due to differences between the test sets, we separately described the results of three subsets of the test (Figure 1).

Test Set 1 consisted of 9 full brain CTA volumes covering the brain upwards from the foramen magnum to evaluate vessel segmentation performance in the entire brain [2]. Test Set 2 consists of 20 ROIs with the same properties as the validation set but from 20 different patients. Lastly, we used a test set (Set 3) of 109 patients with large artery segmentations to evaluate the segmentation of large arteries and specifically the segmentation of the internal carotid artery (ICA) [28]. All patients in Set 3 had an occlusion of the M1 segment of the middle cerebral artery (MCA). Table 1 contains an overview of clinical characteristics and scanner settings for all patients (n=1066) and scans included in this study. Appendix Appendix A contains further details per dataset.

**Table 1:** Dataset characteristics. NIHSS: Severity of stroke symptoms according to National Institutes of Health Stroke Scale. Collateral score: percentage of collateral filling compared to non-infarcted hemisphere. ASPECTS: Alberta Stroke Program Early CT Score. Other occlusion location: M3 or anterior circulation.

| Variable | Value |
| --- | --- |
| Age in years mean(SD) | 69(15) |
| Minutes from stroke onset to treatment mean(SD) | 200(101) |
| NIHSS mean(SD) | 16(11;20) |
| ASPECTS median(IQR) | 9(8;10) |
| Sex N(%) |  |
| Female | 486 (46.7%) |
| Male | 554 (53.3%) |
| Collateral score N(%) |  |
| 0: 0% | 60 (5.9%) |
| 1: 1-50% | 375 (37.1%) |
| 2: 50-99% | 397 (39.3%) |
| 3: 100% | 179 (17.7%) |
| Occlusion segment N(%) |  |
| ICA | 216 (21.4%) |
| M1 | 598 (59.1%) |
| M2 | 187 (18.5%) |
| Other | 11 (1.1%) |
| Manufacturer scanner N(%) |  |
| General Electric | 17 (1.6%) |
| Philips | 208 (19.5%) |
| Siemens | 289 (27.1%) |
| Toshiba | 112 (10.5%) |
| Unknown or other | 440 (41.3%) |
| Slice thickness N(%) | NCCT/CTA |
| $\leq 0.5mm$ | 141 (15.5%)/234 (22%) |
| $>0.5mm$ and $<1.0mm$ | 332 (36.6%)/535 (50.2%) |
| 1.0mm | 435 (47.9%)/297 (27.9%) |
| Peak potential N(%) | NCCT/CTA |
| $\leq 80$ kVp | 11 (1.2%)/167 (15.7%) |
| $>80$ and $\leq 100$ kVp | 253 (27.9%)/587 (55.1%) |
| $>100$ and $\leq 120$ kVp | 642 (70.7%)/308 (28.9%) |
| $>120$ kVp | 2 (0.2%)/4 (0.4%) |

### 2.2. Reference segmentation

To segment vessels, we utilize previously annotated vessel centerlines in the validation set, test Set 1, and test Set 2. A trained observer manually segmented all veins and arteries present as centerlines using the brush and line annotation mode in 3D slicer to construct a reference segmentation [29]. Most notably, the internal carotid artery (ICA), sigmoid sinus, and jugular vein at the point of entering the brain past the bone tissue of the skull were included in the reference segmentation. Test set 3 contained only large artery segmentations, including the ICA, basilar artery, CoW, and MCA which have previously been acquired [28]

### 2.3. Data preparation

To enable voxel-wise comparison between generated and real NCCTs, we performed rigid registration between NCCT and CTA in the training set using Elastix [30]. The registration was performed using normalized cross-correlation as metric, six resolutions with each resolution half the size of the previous or original image, a maximum of 1500 iterations, a maximum step length of 1.0, 2048 random spatial samples, and a Bspline interpolator with degree three. After automated registration, a visual check was performed to rule out severe alignment errors that required another registration attempt. Since deep learning-based image-to-image translation should be largely robust to suboptimal registration in a minority of the cases, we did not exclude NCCT-CTA pairs with minor alignment errors. To standardize the image resolution across the data, we resampled CTAs and NCCTs to a 0.45mm isotropic voxel spacing. This isotropic spacing corresponds to the average spacing in the validation and test sets, which were already preprocessed [2]. Clipping operations were used to minimize and maximize voxel values below and above a threshold. Dual channel CTA images were created by clipping ranges between (0,500) and (500,1000) to distinguish contrast and bone in the scan. The NCCTs were clipped to a (0,500) range to correspond to the CTA. CTA channels and NCCTs were normalized to a (−1,1) range.

## 3. Methods

Contrast fluid in CTA allows to distinguish the vessels from the surrounding tissue in the image which can be used to visualize and segment vessels. Unlike CTA, NCCT does not contain contrast fluid and only visualizes dense structures such as bone and cartilage but not vessels. Hence, the difference between NCCT and CTA mainly comprises contrast fluid in the vessels. We propose to exploit this difference to train an unsupervised method using a GAN to segment vessels in CTA. Furthermore, we add a Frangi-filtering-based loss function to the GAN to emphasize the removal of contrast with tubular structures. The GAN that besides standard GAN losses additionally uses Frangi-loss, is optimized to translate CTAs to NCCTs while at inference only the CTA is used to obtain a vessel segmentation. Thus, reference segmentation is not used during training at any stage. Figure 2 illustrates our approach.

**Figure 2:**
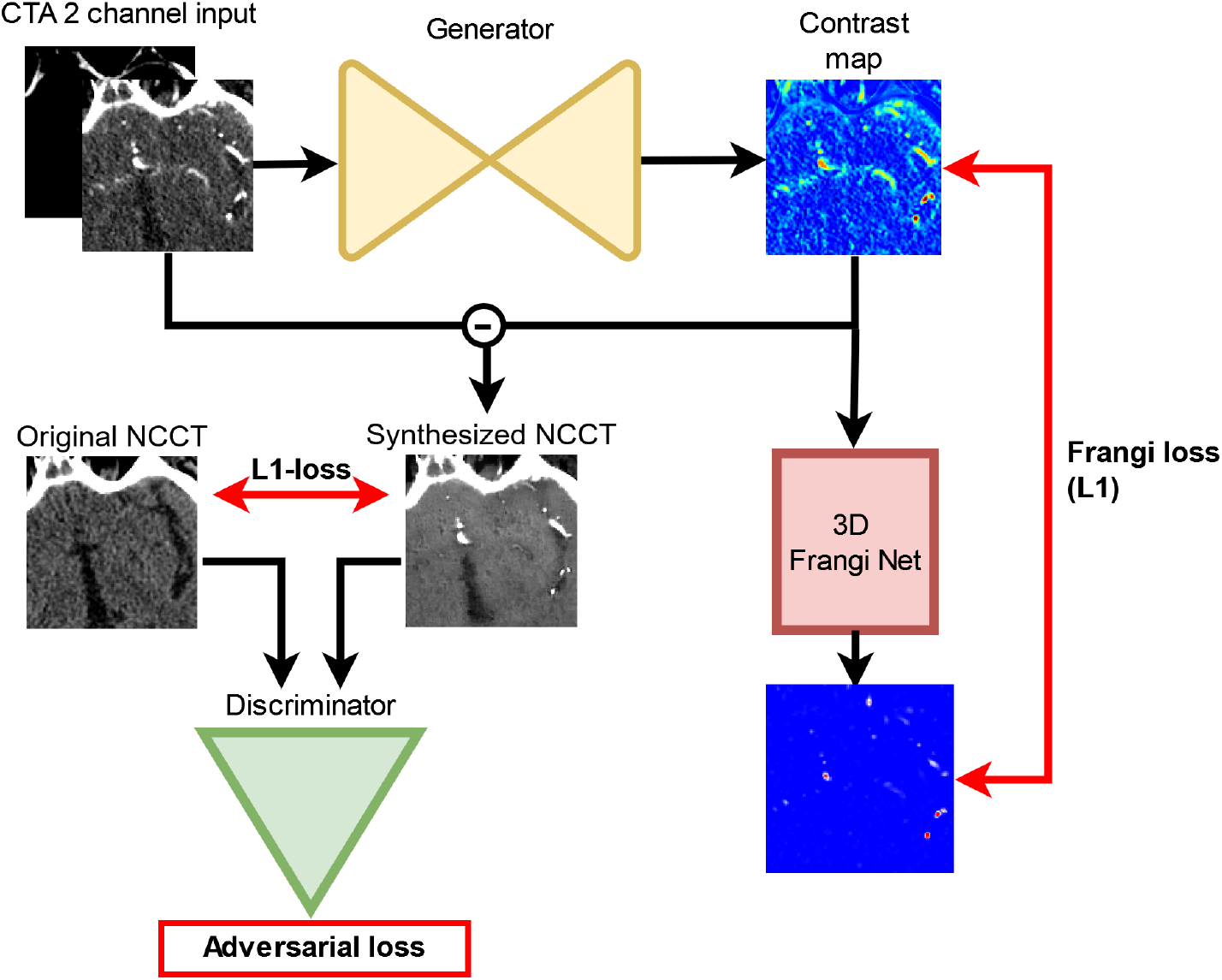
CTA to NCCT translation GAN with Frangi-loss. CTAs are converted to two channel (0,500 and 500,1000) inputs for the generator. The generator synthesizes a contrast map that is subtracted from the input CTA to synthesize an NCCT. The synthesized NCCT is compared with the real co-registered NCCT using the L1-loss and adversarial loss derived from the discriminator’s classification. A Frangi loss is computed based on the Frangi-filtered version of the contrast map to enhance tubular structures. In red the loss functions are depicted. This GAN approach is conditional as the discriminator evaluates the synthesized NCCT as an instance of the real NCCT pair by stacking the two images channel-wise beside the L1-loss.

### 3.1. CTA to NCCT translation with a GAN

We propose to synthesize a contrast map, the difference in contrast between CTA and NCCT, using a GAN. Vessel segmentations are obtained during inference using only the CTA as input. We use a conditional GAN to enforce similarity between the (NCCT_syn_) and (NCCT_real_) during training. Specifically, a conditional GAN can exploit voxelwise similarities between the input (CTA) and output (NCCT) image types since the CTA and (NCCT_real_) visualize the same structures with and without contrast. Furthermore, the similarity in neurovascular structures across the CTA-NCCT pair is maintained using a conditional GAN. The generator (G) of the GAN synthesizes an image containing contrast fluid from a CTA (*Cmap* = *G*(*CTA*)). We refer to this image as the contrast map (Cmap), representing all contrast present in a CTA that should not be present in the corresponding NCCT. The Cmap is subtracted from the CTA to synthesize the NCCT (*NCCT* _syn_ = *CTA* − *Cmap*). The conditional GAN combines a voxel-wise L1 loss between NCCT_syn_ and NCCT_real_ with the adversarial loss (Adv) derived from the discriminator (D). The generator CNN is optimized using the combination of loss functions (Equation 1) consisting of L1 (Equation 2), adversarial (Equation 3), and Frangi-loss (Equation 4). The discriminator distinguishes NCCT_syn_ and NCCT_real_ using a binary cross-entropy loss for the predicted classifications with a label of value 1 (Equation 5). During training, the discriminator is optimized to classify NCCT_syn_ and NCCT_real_ based on a binary cross-entropy loss using labels 0 and 1 respectively.

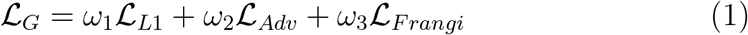

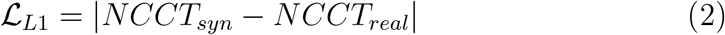

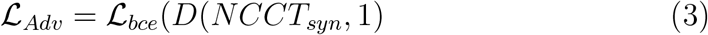

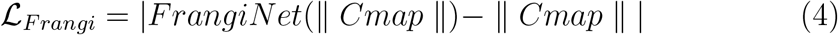

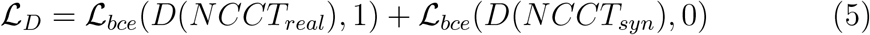

Ideally, the contrast map would only include vessels. However, differences in scan acquisition, processing, and alignment between CTA and NCCT might result in artifacts in the contrast map and the generated vessel segmentation. Furthermore, cartilage and calcifications could result in contrast map artifacts. Compared to the vessels shown in the contrast map, these artifacts are unlikely to have a tubular shape. A Frangi-filter operation on the contrast map can be used to estimate how tubular or vessel-like structures in the contrast map are. Instead of applying a Frangi-filter to the contrast map to obtain a segmentation of the vessels, we propose to use it during training as a loss function to provide feedback to the generator. Thus, the generator will consider the degree of vesselness structures when generating a contrast map.

To compute the Frangi-loss, we propose the 3D FrangiNet in line with a previous 2D version developed by Fu et al. [31]. We defined the mean absolute contrast between the normalized contrast map and the Frangi-filtered normalized contrast map as the Frangi-loss (Fr) (equation 4). Frangi-filtering is performed with the 3D FrangiNet. Subsequently, the gradients of the Frangi-loss are backpropagated through the 3D FrangiNet to optimize the generator. Since vessels smaller than the venous sinuses, ICA, and MCA have a diameter *<* 2.5*mm*, we used predefined *σ* values of 1 and 2 voxels (0.9-1.8mm) to optimize the Hessian filter for these diameters [32]. During training, the weights of the FrangiNet are fixed. Hence, the network is used as a filter to compute the Frangi loss. In Appendix Appendix B, details of the Frangi Net implementation are given.

### 3.2. Generator and discriminator architectures

Figure 3 illustrates the GAN’s generator and discriminator architectures. The generator is based on a 3D Unet with four down and up convolution steps halving and doubling the spatial dimension of the input, respectively. The number of down convolution steps halving the spatial dimension (Gn-down=4, Dn-down=3) and the number of parameters (Gn-features ≈7.1 million, Dn-features ≈2.1 million) for both the generator and discriminator were balanced following Isola et al. to enable stable optimization and prevent mode collapse [33]. Before each down and up convolution, a block with a skip convolution connection and two same convolution layers is placed. In the same convolutions, the number of convolution filters is doubled while the spatial dimension is maintained. The skip convolution connection adds the input of the first same convolution to the output of the second same convolution after doubling the number of features and applying instance normalization. After each second same convolution, a skip connection with the up-convolution part of the Unet is made by copying and concatenating the feature maps.

**Figure 3:**
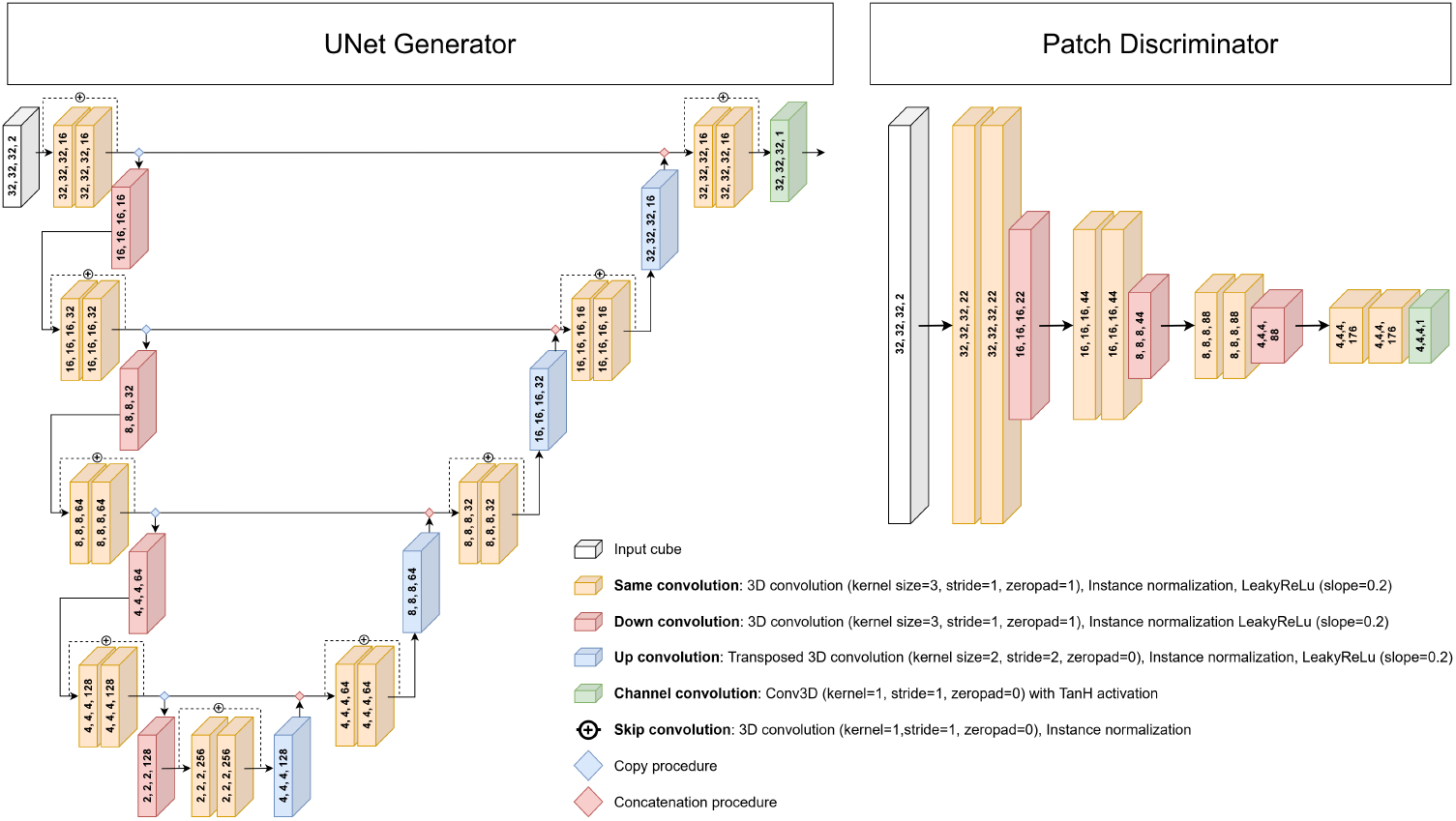
Generator and Discriminator architectures.

The discriminator is based on a 3D patch generator with three down convolutions preceded by two same convolutions without skip convolution. For both the generator and discriminator, all up, down, and same convolution layers are followed by an instance normalization layer and leaky ReLu layer with a slope of 0.2. A Hyperbolic Tangent (TanH) activation function is used as an output layer for both the generator and discriminator.

### 3.3. Postprocessing

To obtain a vessel segmentation at inference, we only use the generator to synthesize contrast maps from CTAs. Since the contrast maps contain degrees of contrast detected by the GAN, a thresholded version of this contrast map is required to obtain a vessel segmentation. The threshold is optimized using the validation set.

As the output from the generator, the contrast map, does not cover a full CTA volume, we sample CTA cubes with the required input size with a stride half the size of the input using Torch.io’s grid sampler [34]. Overlapping areas are aggregated using Gaussian weights over the sampled patches to reduce segmentation errors close to the boundaries of the input cubes. Subsequently, to remove the outer parts of the skull and retain the vascular structures on the inner side of the skull, we remove false positive segmentations outside a 3 mm dilated brain mask previously described by Su et al. [2].

### 3.4. Evaluation

We report performance metrics across volumes in each dataset as median with interquartile range (IQR). Vessel segmentation algorithms can have multiple uses depending on their performance for large or small vessels and high and low contrast attenuation in vessels. Therefore, we report vessel type-specific evaluation metrics considering the diameter, contrast attenuation, and location of vessels. Lastly, we evaluate the false positive rate close to the skull base as this is a common area of segmentation errors [18].

We evaluate the voxel-wise performance per volume using the Dice similarity coefficient (DSC), true positive rate (TPR), and false positive rate (FPR). Although these metrics give a good performance overview across the evaluated volumes, they do not characterize specific errors. False negatives of small vessels contribute less to TPR and DSC than true positives of larger vessels. Therefore, we additionally evaluated the vessel-subtype specific TPR.

To compute vessel-subtype specific metrics, we first separate the reference segmentations into subsegments separated by bifurcations using the VesselVio framework [35]. For each subsegment, we compute the average diameter along the centerline and the average attenuation to separate vessels with a small average diameter (*<* 2.5*mm*), a large average diameter (≥ 2.5*mm*), a low average attenuation (*<* 100*HU*), and a high average attenuation (≥ 100*HU*).

To enable comparison between volumes with varying numbers of vessels, we first compute the median voxelwise TPR for each vessel subtype in a volume, considering all true positive segmentations of other vessel subtypes as true negatives. We report the median and IQR of median TPRs per vessel subtype across all volumes.

False positive segmentations are common close to the base of the skull where the ICA originates. Therefore, we report the FPR in a 10mm radius surrounding the ICA reference segmentations besides the TPR of both ICAs per volume.

We only reported vessel-subtype performance metrics for Test Sets 1 and 2 as Test Set 2 only considers 128×128×128 sized ROIs from different locations of the brain. These ROIs did not have representative distributions of vessel diameter and attenuation to the CTA volumes. We did not perform statistical tests to compare approaches due to small sample sizes and heterogeneity of datasets.

## 4. Experiments and results

First, we describe the fine-tuning of the training parameters we iteratively performed. Subsequently, we report the effect on vessel segmentation performance of each separate loss function or combined loss functions to optimize the generator. Lastly, we compare our approach with a state-off-the-art supervised nnUnet and an other unsupervised approach.

### 4.1. Experimental settings

For the optimization of the discriminator and generator, we used two Adam optimizers with a learning rate of 0.00002 and a batch size of 2 [36]. Each epoch was defined as one sampled input 3D CTA and NCCT cube per patient from the training set. Training was performed for 1000 epochs, the learning rate was kept equal for the first 500 epochs and subsequently linearly decreased to zero for the next 500 epochs. The weights for the loss functions for the GAN loss were based on Isola et al. [33], *ω*_1_=100 and *ω*_2_=1, and for the Frangi loss empirically defined *ω*_3_=1. We experimented with variations in the loss weights but found only limited performance differences. Label smoothing was applied to the NCCT_real_ images’ class label to prevent mode collapse due to overconfidence in classifications of the discriminator (6); a random value between 0.9 and 1.0 was sampled as the class label. We validated every 10th epoch by applying varying thresholds to the contrast map to evaluate the vessel segmentation. Every validation step thresholds between 0.1 (50 HU) and 0.4 (200 HU) with increments of 0.01 (5 HU) were considered. The optimal threshold was based on the Dice similarity coefficient (DSC) relative to the reference vessel segmentation determined on the validation set. During training, structural and contrast augmentations were applied jointly to the NCCT_syn_, NCCT_real_, and the CTA to represent realistic variations present in the data. We added an unsharp noise mask based on the NCCT_real_ and random Gaussian noise to the NCCT_syn_ to prevent overconfident classifications of generated images by the discriminator which might result in mode collapse. The probability of augmentations applied to NCCT_syn_ was linearly reduced to zero after the 500th epoch. NCCT_real_and CTA were jointly augmented to represent realistic variations in the data: Horizontal flip, random rotation with −20 to +20 degrees, zooming between 0.8 (−20%) and 1.2 (+20%), applying an unsharp noise mask, Gaussian filtering to blur the images, adding a random value between −20 and 20 HU to the input images, adding beam hardening noise as previously described (35).

Separate NCCT_real_ augmentations were used to account for contrast differences between CTA and NCCT: adding a random value between 0 and 20, and multiplying the intensities by a value between 0.8 and 1.2. All structural augmentations occurred with a probability of 0.5 per epoch. All contrast augmentations occurred with a probability of 0.2 per epoch.

Since contrast maps generated by the generator only process cubes of a volume, we used Torch.io’s gridsampler to reconstruct contrast maps representing entire CTA volumes [34]. We sample CTA cubes using a stride half the size of the cube to serve as input for the generator. Overlapping voxels in the sampled cubes are aggregated by taking a weighted average. The weights of each voxel a cube are based on Gaussian function with the middle of the cube as peak. Lastly, in line with previous research [2], a 3mm dilated brain mask is used to remove false positive outside of the brain.

### 4.2. Results

Table 2 lists results of our vessel segmentation for each test set separately. To evaluate the benefit of increasing the receptive field for our method (L1+Adv+Fr), we varied input sizes of 32×32×32 (ours (L1+Adv+Fr-32)) to 64×64×64 (ours (L1+Adv+Fr-64)) voxels, as shown in Table 2. The results show that vessel segmentations have similar DSC and TPR values for both input sizes in Test Set 1. However, the FPR was lower for the larger sized input. In test Set 2 the smaller input resulted in better results for all performance metrics. Qualitative evaluation depicted in Figures 4 and 5, revealed that most false positive segmentations of our method are close to the ICA and skull base. Furthermore, scans with higher noise levels also had more false positive segmentations.

**Table 2:**
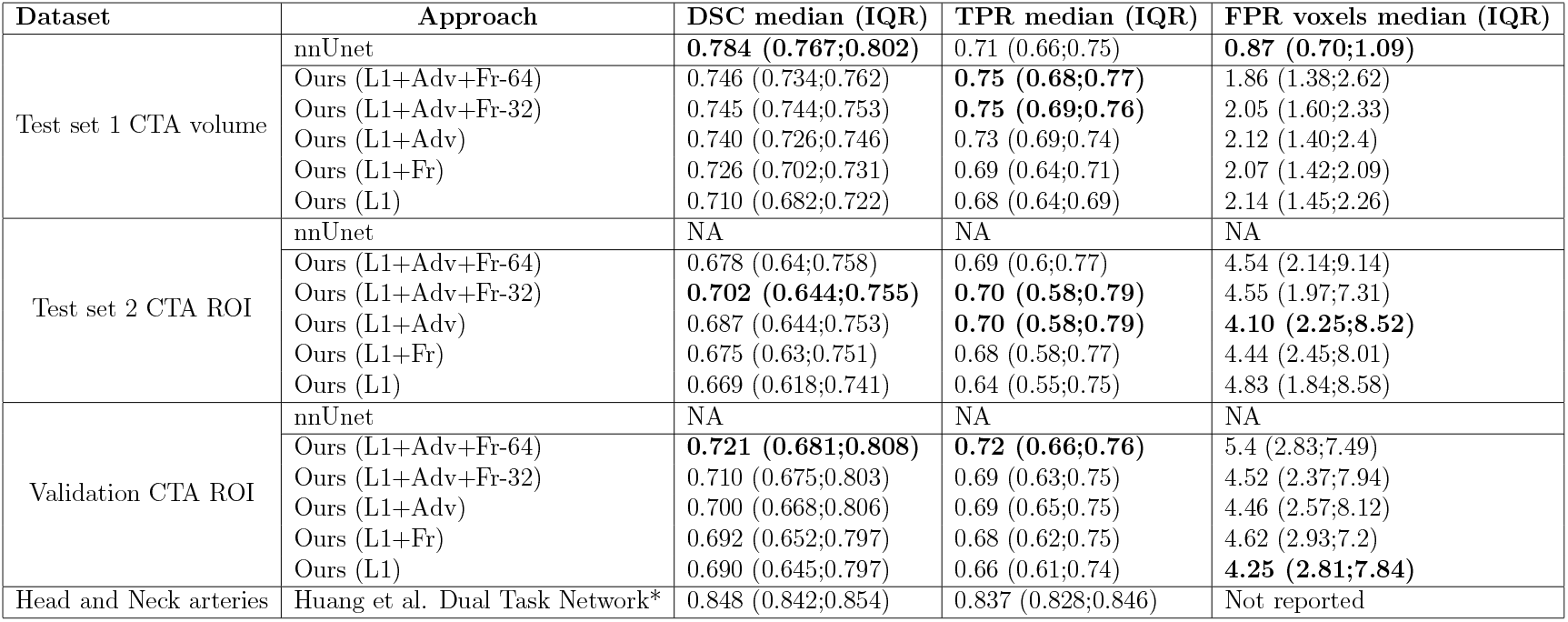
Voxel-wise evaluation metrics. Values are represented as median with interquartile range (IQR). FPR is reported per 1000 voxels. Ours (L1+adv+Fr-64) and (L1+adv+Fr-32): A conditional GAN with Frangi-loss was trained using inputs sized 64×64×64 and 32×32×32 voxels, respectively. Results were also reported for models optimized with loss functions considering the conditional GAN (L1+Adv), L1 and Frangi-loss combines (L1+Fr), and only the L1 loss (L1). *:Huang et al. trained reported performance only for arteries and considered large arteries in the neck for a private single-center dataset, complicating a direct comparison to our study. NA: nnUnet was only evaluated for the Test Set 1 as the other datasets did not include large enough input ROIs.

| Dataset | Approach | DSC median (IQR) | TPR median (IQR) | FPR voxels median (IQR) |
| --- | --- | --- | --- | --- |
| Test set 1 CTA volume | nnUnet | **0.784 (0.767;0.802)** | 0.71 (0.66;0.75) | **0.87 (0.70;1.09)** |
|  | Ours (L1+Adv+Fr-64) | 0.746 (0.734;0.762) | **0.75 (0.68;0.77)** | 1.86 (1.38;2.62) |
|  | Ours (L1+Adv+Fr-32) | 0.745 (0.744;0.753) | **0.75 (0.69;0.76)** | 2.05 (1.60;2.33) |
|  | Ours (L1+Adv) | 0.740 (0.726;0.746) | 0.73 (0.69;0.74) | 2.12 (1.40;2.4) |
|  | Ours (L1+Fr) | 0.726 (0.702;0.731) | 0.69 (0.64;0.71) | 2.07 (1.42;2.09) |
|  | Ours (L1) | 0.710 (0.682;0.722) | 0.68 (0.64;0.69) | 2.14 (1.45;2.26) |
| Test set 2 CTA ROI | nnUnet | NA | NA | NA |
|  | Ours (L1+Adv+Fr-64) | 0.678 (0.64;0.758) | 0.69 (0.6;0.77) | 4.54 (2.14;9.14) |
|  | Ours (L1+Adv+Fr-32) | **0.702 (0.644;0.755)** | **0.70 (0.58;0.79)** | 4.55 (1.97;7.31) |
|  | Ours (L1+Adv) | 0.687 (0.644;0.753) | **0.70 (0.58;0.79)** | **4.10 (2.25;8.52)** |
|  | Ours (L1+Fr) | 0.675 (0.63;0.751) | 0.68 (0.58;0.77) | 4.44 (2.45;8.01) |
|  | Ours (L1) | 0.669 (0.618;0.741) | 0.64 (0.55;0.75) | 4.83 (1.84;8.58) |
| Validation CTA ROI | nnUnet | NA | NA | NA |
|  | Ours (L1+Adv+Fr-64) | **0.721 (0.681;0.808)** | **0.72 (0.66;0.76)** | 5.4 (2.83;7.49) |
|  | Ours (L1+Adv+Fr-32) | 0.710 (0.675;0.803) | 0.69 (0.63;0.75) | 4.52 (2.37;7.94) |
|  | Ours (L1+Adv) | 0.700 (0.668;0.806) | 0.69 (0.65;0.75) | 4.46 (2.57;8.12) |
|  | Ours (L1+Fr) | 0.692 (0.652;0.797) | 0.68 (0.62;0.75) | 4.62 (2.93;7.2) |
|  | Ours (L1) | 0.690 (0.645;0.797) | 0.66 (0.61;0.74) | **4.25 (2.81;7.84)** |
| Head and Neck arteries | Huang et al. Dual Task Network* | 0.848 (0.842;0.854) | 0.837 (0.828;0.846) | Not reported |

**Figure 4:**
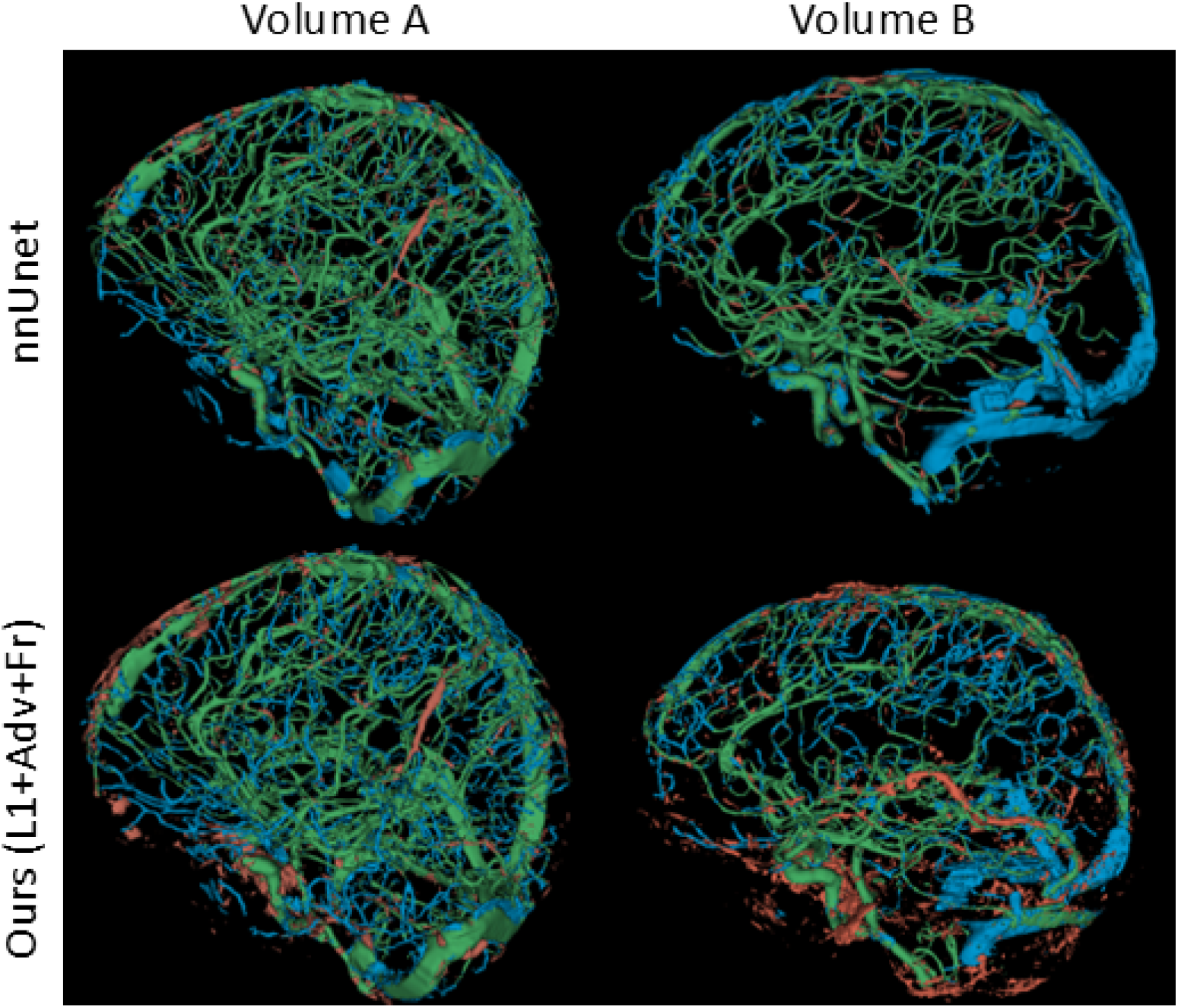
Rendering of automated segmentation quality compared to an expert-based reference segmentation. Column 1 (Volume A): CTA with low noise level. Column 2 (Volume B): CTA with high noise level. Row 1 shows the segmentation performance of the state-off-the-art nnUnet. Row 2 shows the performance of our unsupervised method (L1+Fr+Adv-32). True positives are shown in green, false positives in brown, false negatives in blue.

**Figure 5:**
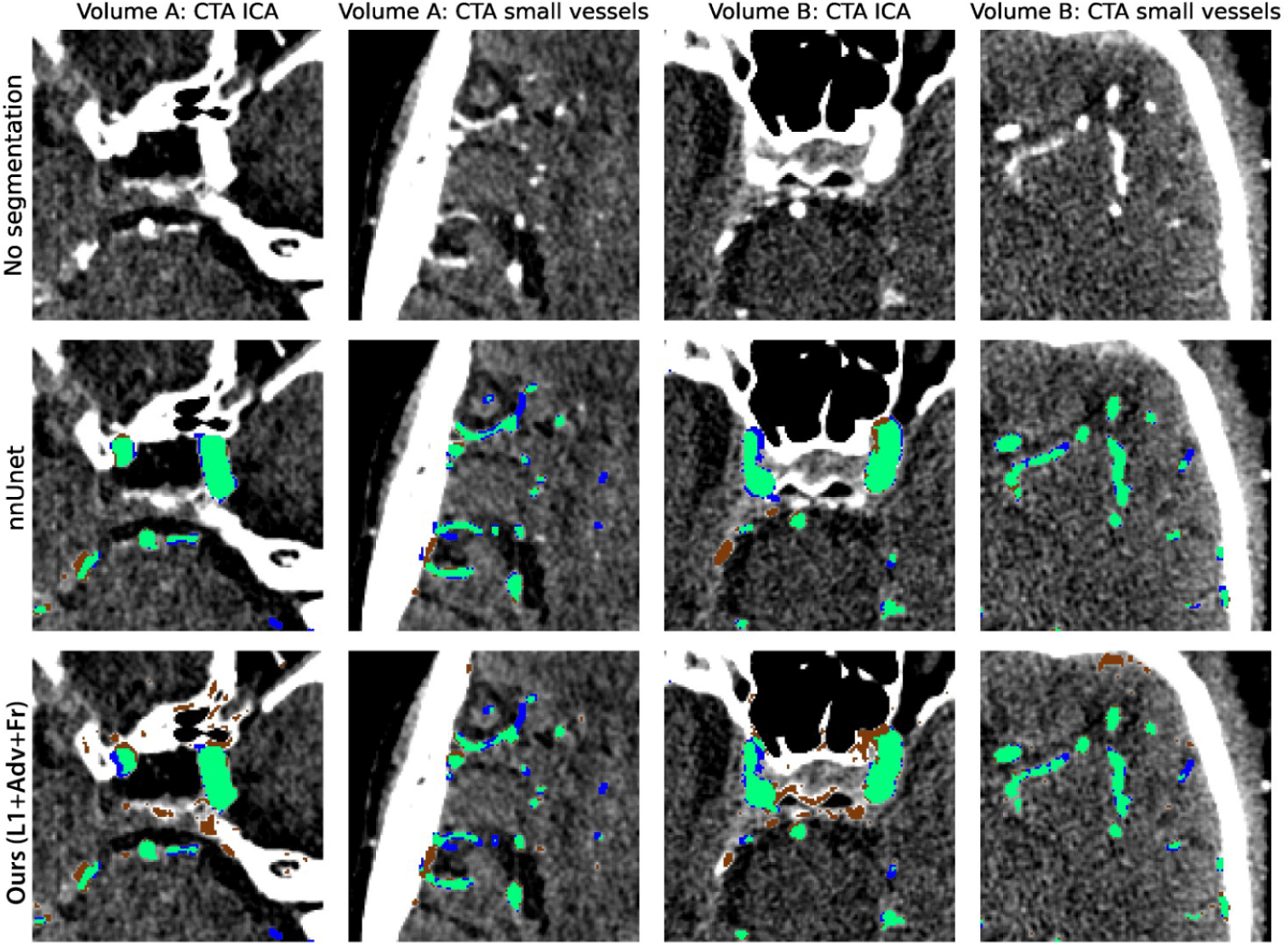
Cross section of automated segmentation results (same cases as Figure 4 - Row: 1) CTA without segmentations, 2) segmentation of the nnUnet, 3) segmentation of our unsupervised method (L1+Fr+Adv-32). Columns: For Volume A with low noise levels (column 1 and 2) and Volume B with higher noise levels (column 2 and 3) automatic segmentation performance close to the ICA (columns 1 and 3) and more cranial in the brain for small diameter vessels (columns 2 and 4). True positives are shown in green, false positives in brown, false negatives in blue.

Performance of our method was evaluated separately for vessels of small (*<* 2.5*mm*) and large (≥ 2.5*mm*) diameter, as well as for the vessels of lower (*<* 100*HU*) and higher(≥ 100*HU*) attenuation. The results are presented in Table 3; the results show that TPR was higher for large-diameter vessels. Furthermore, for both Test Sets 1 and 2, the TPR was higher for vessels with an average attenuation ≥ 100 HU compared to vessels with an average attenuation *<* 100*HU*. From qualitative evaluation (Figure 4 and 5) and quantitative comparison of FPRs between the main volume-wise results (Table 2) and the 10 mm area surrounding the ICA (Table 4), it appears that false positive segmentations were present primarily close to the skull base and surrounding the ICAs.

**Table 3:** True positive rates per vessel-subtype. The results are separately shown for vessels with a small (*<* 2.5*mm*) and large (≥ 2.5*mm*) diameter, and for vessels of low (*<* 100*HU*) and high(≥ 100*HU*) contrast attenuation. The median and IQR TPR per volume are computed after selecting only a specific subset of vessels in each volume (columns). TPRs are presented as median and interquartile range (IQR) across median TPRs per volume. This TPR considers all other segmented vessel subtypes as true negatives. Performance of supervised (nnUnet), and our unsupervised method trained with 32×32×32 voxel-sized inputs are presented: combining L1, adversarial, and Frangi loss (L1+Adv+Fr), conditional GAN (L1+Adv), L1 and Frangi-loss combined (L1+Fr), and only the L1 loss (L1).

| Dataset | Approach | Vessel subtype: Average vessel diameter |  | Vessel subtype: Average vessel attenuation |  | ICA |
| --- | --- | --- | --- | --- | --- | --- |
|  |  | <2.5mm | ≥2.5mm | <100 HU | ≥100 HU |  |
| Test Set 1 CTA volume | nnUnet | **0.65 (0.61;0.75)** | 0.87 (0.77;0.92) | **0.46 (0.41;0.53)** | 0.80 (0.71;0.85) | 0.79 (0.76;0.84) |
|  | Ours (L1+Adv+Fr-32) | 0.62 (0.52;0.65) | **0.88 (0.87;0.89)** | 0.30 (0.27;0.36) | **0.84 (0.82;0.86)** | 0.89 (0.76;0.92) |
|  | Ours (L1+Adv) | 0.62 (0.49;0.66) | 0.87 (0.85;0.88) | 0.29 (0.25;0.34) | 0.84 (0.81;0.85) | **0.89 (0.85;0.92)** |
|  | Ours (L1+Fr) | 0.60 (0.46;0.65) | 0.83 (0.80;0.84) | 0.28 (0.26;0.32) | 0.80 (0.77;0.83) | 0.68 (0.6;0.76) |
|  | Ours (L1) | 0.59 (0.46;0.63) | 0.81 (0.77;0.82) | 0.26 (0.25;0.32) | 0.79 (0.75;0.81) | 0.67 (0.65;0.77) |
| Test Set 3 Large arteries | nnUnet | 0.95 (0.89;0.99) | **0.98 (0.96;1.0)** | **0.79 (0.69;0.86)** | 0.95 (0.91;0.99) | **0.91 (0.80;0.96)** |
|  | Ours (L1+Adv+Fr-32) | 0.97 (0.90;0.99) | 0.98 (0.93;1.0) | 0.50 (0.43;0.61) | 0.96 (0.92;0.99) | **0.91 (0.80;0.96)** |
|  | Ours (L1+Adv) | **0.97 (0.92;0.99)** | 0.98 (0.92;1.0) | 0.53 (0.44;0.62) | **0.97 (0.93;0.99)** | 0.89 (0.74;0.95) |
|  | Ours (L1+Fr) | 0.97 (0.91;1.0) | 0.97 (0.92;1.0) | 0.51 (0.45;0.63) | 0.96 (0.91;0.99) | 0.87 (0.7;0.94) |
|  | Ours (L1) | 0.95 (0.89;0.99) | 0.98 (0.92;1.0) | 0.50 (0.41;0.60) | 0.94 (0.89;0.98) | 0.87 (0.73;0.94) |

**Table 4:** False positive rate in 10-millimeter radius surrounding the ICA.

| Approach | Test Set 1<br>CTA volume<br>ICA-FPR | Test Set 3<br>Large arteries<br>ICA-FPR |
| --- | --- | --- |
| nnUnet | **0.10 (0.06;0.28)** | **0.21 (0.16;0.28)** |
| Ours (L1+Adv+Fr-32) | 0.84 (0.19;1.08) | 0.72 (0.52;0.97) |
| Ours (L1+Adv) | 0.82 (0.21;1.13) | 0.71 (0.55;0.92) |
| Ours (L1+Fr) | 0.68 (0.21;0.95) | 0.68 (0.5;0.88) |
| Ours (L1) | 0.85 (0.17;1.11) | 0.69 (0.51;0.87) |

### 4.3. Contribution of unsupervised losses

To evaluate the relative contribution to performance gains of unsupervised loss functions, we conducted separate experiments using only the L1 loss, L1 and Frangi-loss (L1+Fr), and the conditional GAN combining L1 and Adversarial loss (L1+Adv). Cubes sized 32×32×32 voxels were used as input for the models. Combining the L1 with the Frangi or adversarial loss improved the DSC compared to using the L1 loss alone. Adding the adversarial loss slightly improved the TPR. Adding the Frangi loss resulted in a lower FPR. These results are listed in Table 2. Adding either the Frangi loss or adversarial loss to the L1 improved the median TPR regardless of the vessel’s diameter or contrast attenuation. This improvement was more profound when the adversarial loss was added to the L1.

### 4.4. Comparison with other methods

Our unsupervised method was compared with a state-of-the-art supervised deep learning method for segmentation, nnUnet [37], and to another previously published unsupervised brain vessel segmentation method described by Huang et al. [18].

The nnUnet was optimized in a 9-fold nested cross-validation setting using six volumes for training, two for validation, and one for testing. The nnUnet full-resolution compiler used an input cube size of 160×112×128 and 5-4-5 down-sampling convolutions for each of the three dimensions. Due to the required input of the nnUnet, we did not report evaluation metrics for the 128×128×128 ROI-sized test Set 2.

Huang et al. used a cGAN with an additional self-supervised cross-entropy loss based on self-generated vessel segmentations. These vessel segmentations are acquired by applying an adaptive threshold to the contrast map that results from subtracting the NCCT from the CTA. Subsequently, noise removal and clustering techniques are used to refine these self-generated reference segmentations. Given that the source code and the data used by Huang et al. [18] are not available, we report the performance from their manuscript. We compared the reported metrics and converted them to our results format (median with IQR) described in Table 2. Hence, the numbers are not directly comparable and can only serve as an indication.

Our unsupervised method approached the performance of the supervised nnUnet regarding DSC. However, the nnUnet resulted in a lower FPR and TPR compared to our unsupervised method, suggesting that the nnUnet tended to under-segment vessels compared to our method. In contrast, our method tended to over-segment vessels compared to the nnUnet.

The results show that Huang et al. achieved higher TPR and DSC than our method and than the state-off-the-art nnUnet. Note, however, that Huang et al. presented results for head and neck arteries together, whereas we evaluated arteries and veins in the head.

Figures 4 and 5 illustrate the true positive, false positive, and false negative predicted segmentations of our unsupervised method and the supervised nnUnet for a CTA with a low noise level and high DSC (Volume A; DSC L1+Adv+Fr: 0.75, nnUnet: 0.80) and a CTA with a high noise level and poor DSC (Volume B; DSC L1+Adv+Fr: 0.57, nnUnet: 0.61). Compared to the nnUnet, our unsupervised method had a high degree of false positive segmentations in the skull and cartilage tissue close to the location where the ICAs and Transverse Sinuses enter the skull. Furthermore, our unsupervised method had more false positives if the noise level in the CTA was higher. The nnUnet had worse performance in the venous sinuses. Detailed results are listed in Table 2 and Table 3.

## 5. Discussion

We proposed an unsupervised deep learning method for vessel segmentation in CTA. The method used a conditional GAN for CTA to NCCT translation that in addition to standardly used losses also exploits a Frangi-loss. Instead of direct CTA to NCCT translation using the generator of the conditional GAN, we generated a contrast map. After subtracting this contrast map from the CTA, a synthetic NCCT is generated. This approach minimizes the opportunity for the GAN to change the anatomy substantially, allows us to exploit the properties of vessel-like structures, and is more resistant to noise in CTAs. During training, the L1 and adversarial loss are used to optimize the generator for synthesizing contrast maps representing all the contrast present in input CTA. We introduced an additional 3D Frangi-loss to enforce the removal of contrast with vessel-like shapes. Our unsupervised method achieves state-of-the-art performance for segmenting vessels with a large diameter and high attenuation, such as venous sinuses, ICAs, MCAs, and basilar arteries. However, the segmentation of vessels with a small diameter and poor attenuation remains a challenge. False positive segmentations are frequent in dense tissue adjacent to the ICA but are largely absent in the brain.

Experiments in this study show the added value of each part of the combined loss function used for our method (L1, Adv, and Fr). Our results indicate that the L1-loss can be considered as the backbone of our method. Whereas the adversarial loss enhances the detection of vessels (true positive segmentations), the Frangi-loss reduces false positive segmentation of bone and noise structures that do not have vessels-like shapes.

Previous work by Huang et al. showed better results in unsupervised vessel segmentation for a different dataset, employing a combination of L1-loss for CTA to NCCT translation and a supervised loss based on self-generated vessel segmentations [18]. However, the lack of available data, variations in reference segmentation, and dataset-specific image processing complicate the implementation of the described method and comparison to our work. Specifically, Huang et al. did not consider venous sinuses, and small and low attenuated vessels in the reference segmentation. Furthermore, reference segmentation had a strong emphasis on large extracranial neck arteries. In addition, Huang et al. based their reference segmentations on an initial subtraction image, which closely resembles the method for obtaining self-generated vessel segmentations used for optimizing the CNN. In contrast, we considered all vessels cranial to the foramen magnum and performed manual segmentations without subtraction images.

During training, we used a conditional GAN with a Unet as generator due to the limited computational burden. Depending on the input cube size and reconstruction cube overlap, training of our method (L1+Adv+Fr-32) took 500-1000 seconds (50-500 seconds for the L1+Adv approach) per training epoch while the inference took 2-14 seconds per volume using an NVIDIA Titan V (12GB VRAM, 1455 MHz clock speed). Improvements in unsupervised CTA to NCCT translation might be facilitated by diffusion models or contrastive learning approaches at the cost of a higher computational burden [38, 39].

While state-off-the-art supervised vessel segmentation methods generally outperform unsupervised approaches, their requirement for manual segmentations complicates the training with large and diverse datasets. Our results demonstrate the feasibility of brain vessel segmentation using the proposed unsupervised method but also show that a relatively limited training set allows segmentation with similar and in some cases better performance than a state of the art supervised nnUnet. Nevertheless, considering a manual vessel annotation time frequently exceeding a day per CTA volume, a significant amount of time can be saved when using our approach for training a vessel segmentation model. Furthermore, our approach allows for training on larger and more diverse datasets which likely enhances generalizability.

We have shown that incorporating vessel shape characteristics with 3D Frangi-loss can support the training of unsupervised vessel segmentation models. Future research should aim to optimize the predefined Frangi-filtering settings for specific tasks such as ICA or small vessel segmentation. Additional shape constraints can support anatomically consistent and connected vessel segmentations [40].

Further research based on our unsupervised method for vessel segmentation can warrant improvements in vessel segmentation in CTA. Specifically, the combination of unsupervised pre-training and supervised fine-tuning or supervised training on pseudo segmentations based on our unsupervised method can be considered. Furthermore, recently developed foundation models such as the segment anything model (SAM) seem promising but appear to perform poorly on tasks not represented in public datasets such as vessel segmentation [41]. Nevertheless, SAM approaches can be considered to identify regions, such as the skull base, to enhance vessel segmentation models’ training with dedicated loss functions or regional loss weights.

## 6. Conclusion

We presented a method for unsupervised generative deep learning-based intracranial vessel segmentation. Our method performs a segmentation using a generative adversarial network that combines the mean absolute error, the adversarial loss, and a newly introduced 3D Frangi-loss. Results presented in this study suggest that unsupervised generative deep learning models offer the possibility to segment intracranial vessels approaching state-off-the-art performance without the need for laborious manual segmentations.

## Appendix A. Appendix: Data characteristics

Table A.5 describes dataset characteristics for train, validation, and test sets separately.

**Table A.5:**
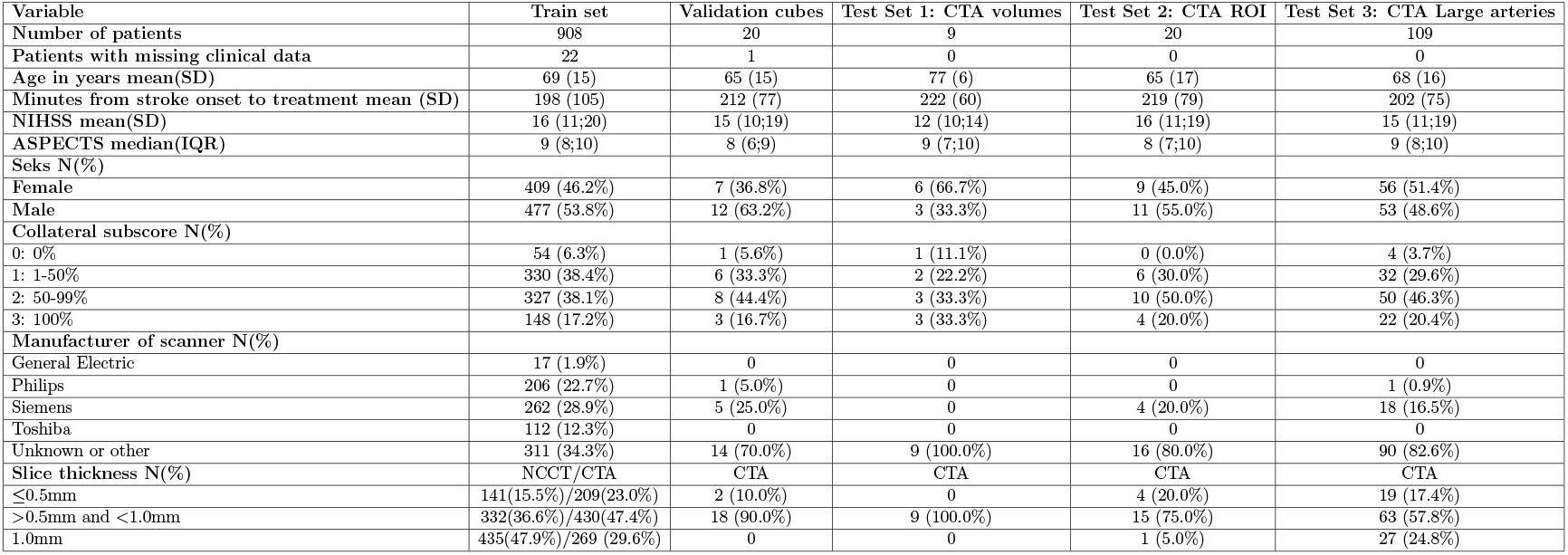
A.5: Dataset characteristics split by train, validation, and test sets

## Appendix B. Appendix: Frangi filter

Details of the implementation of the 3D Frangi Net. First, six 3D convolution filters are used to compute the local Hessian concerning each of the three dimensions. These six convolution filters are based on the second-order derivative (Hessian) concerning each of the three dimensions of a Gaussian kernel. Subsequently, the three eigenvalues (| *λ*_1_| ≤ | *λ*_2_| ≤ | *λ*_3_|) of the Hessian per voxel represent the rate of intensity change in the surrounding voxels.

Based on these eigenvalues the blob-like 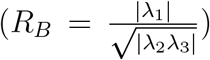, the aspect ratio of the two longest axes 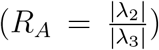,and the second order structuredness 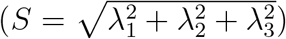 are computed. These measures are combined to compute the vessels measures 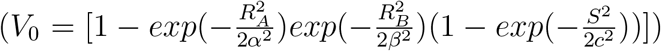. We considered only bright vesselness structures and used predefined values for *α* = 0.5, *β* = 0.5, and *c* = 2 [9]. The standard deviation (*σ*) of the Gaussian kernel used to compute the Hessian results in the highest vesselness measure (*V*_0_) for vessels with a diameter of 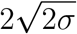 voxels [31, 9]. The maximum *V*_0_ for those two *σ* was used as the final value.

## Appendix C. Appendix: Training examples

In Figure C.6 examples of segmentation differences between methods for a validation set example during various training stages.

**Figure C.6:**
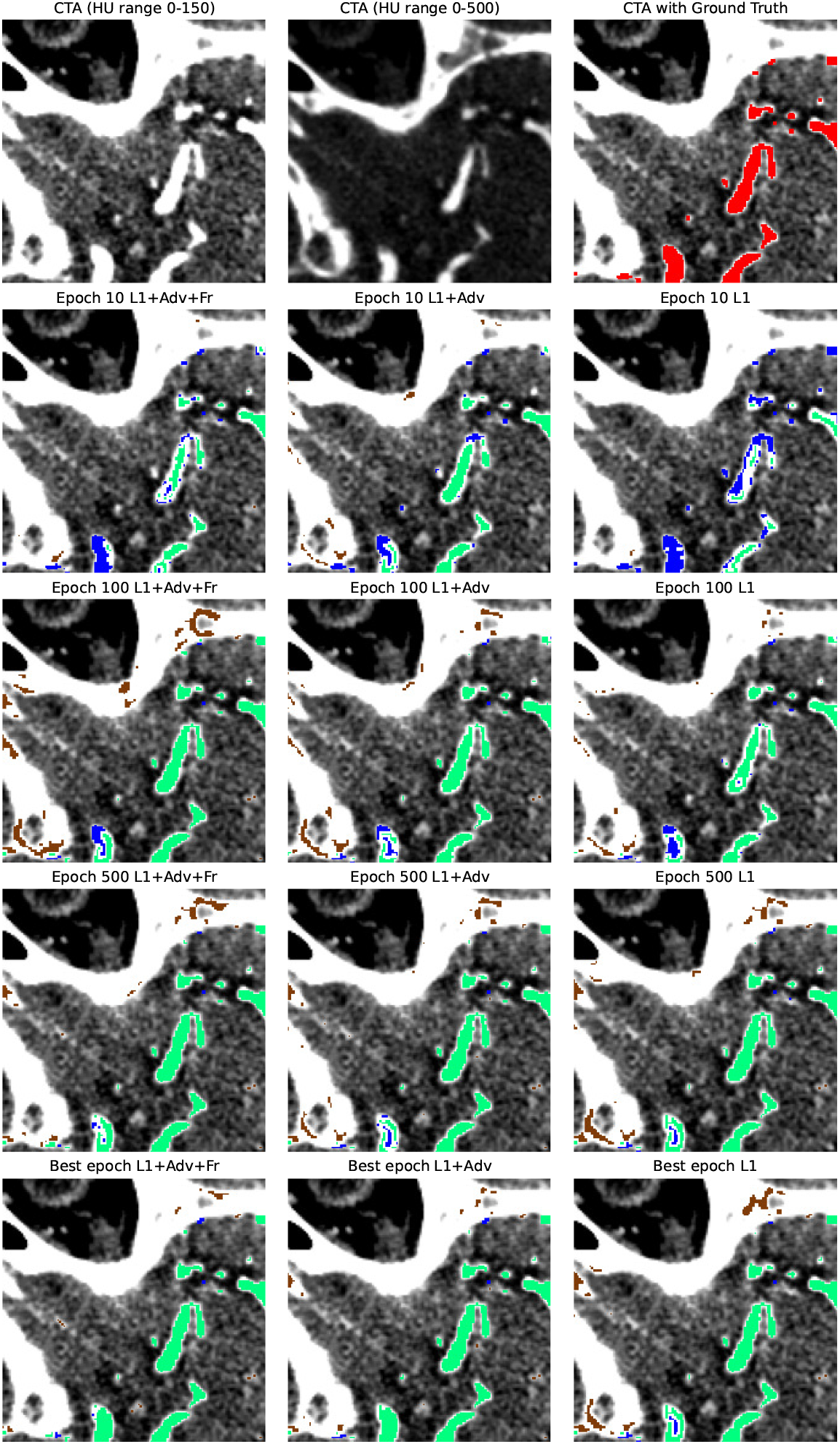
Validation example during training

## Declaration of Generative AI

During the preparation of this work, the author(s) used ChatGPT and Grammarly to adjust written text and correct spelling and style errors. After using this tool/service, the author(s) reviewed and edited the content as needed and take(s) full responsibility for the content of the published article.

## Data availability statement

Data used for this study is available upon reasonable request via an online application form: https://www.contrast-consortium.nl/data-requests-consortium-members-and-trial-collaborators/

## Notes

### Competing Interest Statement

Henk van Voorst reports financial support was provided by Dutch Research Council (NWO-ZonMW). Henk van Voorst reports financial support was provided by Netherlands Heart Foundation. Charles Majoie reports financial support was provided by WIN Foundation, CVON-Dutch Heart Foundation, Stryker, Boehringer Ingelheim, European Commission, Healthcare Evaluation Netherlands, shareholder of Nicolab. Bart Emmer reports financial support was provided by Leading the Change and Dutch Science foundation, from Topsector Life Sciences HealthHolland and Nicolab, all paid to institution. M.W.A. Caan is shareholder of Nico-lab International Ltd.Reports a relationship with that includes:. Has patent pending to. If there are other authors, they declare that they have no known competing financial interests or personal relationships that could have appeared to influence the work reported in this paper.

### Funding Statement

This research was funded by the Dutch Heart Foundation

### Author Declarations

Waiver for IRB approval (ethical committee of Erasmus MC University Medical Centre, Rotterdam, the Netherlands MEC-2014-235) is available due to the retrospective nature of the MR CLEAN registry data.

## References

1. M. T. Stib, J. Vasquez, M. P. Dong, Y. H. Kim, S. S. Subzwari, H. J. Triedman, A. Wang, H. L. C. Wang, A. D. Yao, M. Jayaraman, J. L. Boxerman, C. Eickhoff, U. Cetintemel, G. L. Baird, R. A. McTaggart, Detecting large vessel occlusion at multiphase CT angiography by using a deep convolutional neural network, Radiology 297 (3) (2020) 640–649. doi:10.1148/radiol.2020200334.

2. J. Su, L. Wolff, A. C. Van Es, W. Van Zwam, C. Majoie, D. W. Dippel, A. Van Der Lugt, W. J. Niessen, T. Van Walsum, Automatic Collateral Scoring from 3D CTA Images, IEEE Transactions on Medical Imaging 39 (6) (2020) 2190–2200. doi:10.1109/TMI.2020.2966921.

3. L. Winkelmeier, J. J. Heit, G. Adusumilli, V. Geest, A. Guenego, G. Broocks, J. Prüter, N. O. Gloyer, L. Meyer, H. Kniep, M. G. Lansberg, G. W. Albers, M. Wintermark, J. Fiehler, T. D. Faizy, Poor venous outflow profiles increase the risk of reperfusion hemorrhage after endovascular treatment, Journal of Cerebral Blood Flow and Metabolism (2022). doi:10.1177/0271678×221127089.

4. F. Bukenya, L. Bai, A. Kiweewa, A Review of Blood Vessel Segmentation Techniques, 1st International Conference on Computer Applications and Information Security, ICCAIS 2018 (2018) 1– 10doi:10.1109/CAIS.2018.8441989.

5. S. Moccia, E. De Momi, S. El Hadji, L. S. Mattos, Blood vessel segmentation algorithms — Review of methods, datasets and evaluation metrics, Computer Methods and Programs in Biomedicine 158 (2018) 71–91. doi:10.1016/j.cmpb.2018.02.001. URL https://doi.org/10.1016/j.cmpb.2018.02.001

6. R. Manniesing, M. A. Viergever, W. J. Niessen, Vessel axis tracking using topology constrained surface evolution, IEEE Transactions on Medical Imaging 26 (3) (2007) 309–316.

7. S. Cetin, G. Unal, A higher-order tensor vessel tractography for segmentation of vascular structures, IEEE transactions on medical imaging 34 (10) (2015) 2172–2185.

8. H. Hassanpour, N. Samadiani, S. M. M. Salehi, Using morphological transforms to enhance the contrast of medical images, The Egyptian Journal of Radiology and Nuclear Medicine 46 (2) (2015) 481–489.

9. A. F. Frangi, W. J. Niessen, K. L. Vincken, M. A. Viergever, Multiscale vessel enhancement filtering, Lecture Notes in Computer Science (including subseries Lecture Notes in Artificial Intelligence and Lecture Notes in Bioinformatics) 1496 (February) (1998) 130–137. doi:10.1007/bfb0056195.

10. P. Lu, J. Xia, Z. Li, J. Xiong, J. Yang, S. Zhou, L. Wang, M. Chen, C. Wang, A vessel segmentation method for multi-modality angiographic images based on multi-scale filtering and statistical models, Biomedical engineering online 15 (1) (2016) 1–18.

11. K. T. Bae, Intravenous contrast medium administration and scan timing at CT: considerations and approaches, Radiology 256 (1) (2010) 32–61.

12. O. Ronneberger, P. Fischer, T. Brox, U-net: Convolutional networks for biomedical image segmentation, Lecture Notes in Computer Science (including subseries Lecture Notes in Artificial Intelligence and Lecture Notes in Bioinformatics) 9351 (2015) 234–241. doi:10.1007/978-3-319-24574-428.

13. A. Hilbert, V. I. Madai, E. M. Akay, O. U. Aydin, J. Behland, J. Sobesky, I. Galinovic, A. A. Khalil, A. A. Taha, J. Wuerfel, P. Dusek, T. Niendorf, J. B. Fiebach, D. Frey, M. Livne, BRAVE-NET: Fully Automated Arterial Brain Vessel Segmentation in Patients With Cerebrovascular Disease, Frontiers in Artificial Intelligence 3 (September) (2020) 1–14. doi:10.3389/frai.2020.552258.

14. G. Tetteh, V. Efremov, N. D. Forkert, M. Schneider, J. Kirschke, B. Weber, C. Zimmer, M. Piraud, B. H. Menze, DeepVesselNet: Vessel Segmentation, Centerline Prediction, and Bifurcation Detection in 3-D Angiographic Volumes (2018) 1–13. URL http://arxiv.org/abs/1803.09340

15. S. E. Hadji, S. Moccia, D. Scorza, M. Rizzi, F. Cardinale, G. Baselli, E. D. Momi, Brain-vascular segmentation for SEEG planning via a 3D fully-convolutional neural network, Proceedings of the Annual International Conference of the IEEE Engineering in Medicine and Biology Society, EMBS (2019) 1014–1017doi:10.1109/EMBC.2019.8857456.

16. P. Sanches, C. Meyer, V. Vigon, B. Naegel, Cerebrovascular network segmentation of MRA images with deep learning, arXiv (Isbi) (2018) 768–771.

17. Y. Wang, G. Yan, H. Zhu, S. Buch, Y. Wang, E. M. Haacke, J. Hua, Z. Zhong, VC-Net: Deep Volume-Composition Networks for Segmentation and Visualization of Highly Sparse and Noisy Image Data, IEEE Transactions on Visualization and Computer Graphics (2020) 1– 1doi:10.1109/tvcg.2020.3030374.

18. W. Huang, W. Gao, C. Hou, X. Zhang, X. Wang, J. Zhang, Simultaneous vessel segmentation and unenhanced prediction using self-supervised dual-task learning in 3D CTA (SVSUP), Computer Methods and Programs in Biomedicine 224 (2022) 107001. doi:10.1016/j.cmpb.2022.107001. URL https://doi.org/10.1016/j.cmpb.2022.107001

19. S. Liu, R. Su, J. Su, J. Xin, J. Wu, W. van Zwam, P. J. van Doormaal, A. van der Lugt, W. J. Niessen, N. Zheng, others, An automated framework for brain vessel centerline extraction from CTA images, arXiv preprint 2401.07041 (2024).

20. J. Su, S. Li, L. Wolff, W. van Zwam, W. J. Niessen, A. van der Lugt, T. van Walsum, Deep reinforcement learning for cerebral anterior vessel tree extraction from 3D CTA images, Medical image analysis 84 (2023) 102724.

21. R. L. M. van Herten, I. Lagogiannis, T. Leiner, I. Išgum, The role of artificial intelligence in coronary CT angiography, Netherlands Heart Journal (2024) 1–9.

22. I. Goodfellow, Y. Bengio, A. Courville, Deep learning, MIT Press, 2016. URL deeplearningbook.org

23. I. J. Goodfellow, J. Pouget-Abadie, M. Mirza, B. Xu, D. Warde-Farley, S. Ozair, A. Courville, Y. Bengio, jGenerative adversarial nets, Vol. 3, 2014.

24. S. Xun, D. Li, H. Zhu, M. Chen, J. Wang, J. Li, M. Chen, B. Wu, H. Zhang, X. Chai, Z. Jiang, Y. Zhang, P. Huang, Generative adversarial networks in medical image segmentation: A review, Computers in Biology and Medicine 140 (2022). doi:10.1016/j.compbiomed.2021.105063. URL https://doi.org/10.1016/j.compbiomed.2021.105063

25. Z. Chen, L. Xie, Y. Chen, Q. Zeng, Q. ZhuGe, J. Shen, C. Wen, Y. Feng, Generative adversarial network based cerebrovascular segmentation for time-of-flight magnetic resonance angiography image, Neurocomputing 488 (2022) 657–668.

26. I. G. H. Jansen, M. J. H. L. Mulder, R.-J. B. Goldhoorn, MR CLEAN Registry investigators, Endovascular treatment for acute ischaemic stroke in routine clinical practice: prospective, observational cohort study (MR CLEAN Registry)., BMJ (Clinical research ed.) 360 (2018) k949. doi:10.1136/bmj.k949.

27. W. J. Powers, A. A. Rabinstein, T. Ackerson, O. M. Adeoye, N. C. Bambakidis, K. Becker, J. Biller, M. Brown, B. M. Demaerschalk, B. Hoh, E. C. Jauch, C. S. Kidwell, T. M. Leslie-Mazwi, B. Ovbiagele, P. A. Scott, K. N. Sheth, A. M. Southerland, D. V. Summers, D. L. Tirschwell, Guidelines for the early management of patients with acute ischemic stroke: 2019 update to the 2018 guidelines for the early management of acute ischemic stroke a guideline for healthcare professionals from the American Heart Association/American Stroke A (2019). doi:10.1161/STR.0000000000000211.

28. C. Miller, P. Konduri, S. Bridio, G. Luraghi, N. Arrarte, N. Boodt, N. Samuels, J. F. Rodriguez, F. Migliavacca, H. Lingsma, A. V. D. Lugt, Y. Roos, D. Dippel, H. Marquering, C. Majoie, A. Hoekstra, Computer Methods and Programs in Biomedicine In silico thrombectomy trials for acute ischemic stroke, Computer Methods and Programs in Biomedicine 228 (2023) 107244. doi:10.1016/j.cmpb.2022.107244. URL https://doi.org/10.1016/j.cmpb.2022.107244

29. 3D Slicer image computing platform — 3D Slicer. URL https://www.slicer.org/

30. S. Klein, M. Staring, K. Murphy, M. A. Viergever, J. P. Pluim, Elastix: A toolbox for intensity-based medical image registration, IEEE Transactions on Medical Imaging 29 (1) (2010) 196–205. doi:10.1109/TMI.2009.2035616.

31. W. Fu, K. Breininger, T. Würfl, N. Ravikumar, R. Schaffert, A. Maier, Frangi-Net: A Neural Network Approach to Vessel Segmentation (2017) 1–6. URL http://arxiv.org/abs/1711.03345

32. A. T. Rai, J. P. Hogg, B. Cline, G. Hobbs, Cerebrovascular geometry in the anterior circulation: An analysis of diameter, length and the vessel taper, Journal of NeuroInterventional Surgery 5 (4) (2013) 371–375. doi:10.1136/neurintsurg-2012-010314.

33. P. Isola, J. Y. Zhu, T. Zhou, A. A. Efros, Image-to-image translation with conditional adversarial networks, Proceedings - 30th IEEE Conference on Computer Vision and Pattern Recognition, CVPR 2017 2017-Janua (2017) 5967–5976. doi:10.1109/CVPR.2017.632.

34. F. Pérez-Garcia, R. Sparks, S. Ourselin, TorchIO: a Python library for efficient loading, preprocessing, augmentation and patch-based sampling of medical images in deep learning, Computer Methods and Programs in Biomedicine 208 (2021) 106236.

35. J. R. Bumgarner, R. J. Nelson, Open-source analysis and visualization of segmented vasculature datasets with VesselVio, Cell Reports Methods 2 (4) (2022) 100189. doi:10.1016/j.crmeth.2022.100189. URL https://doi.org/10.1016/j.crmeth.2022.100189

36. D. P. Kingma, J. L. Ba, Adam: A method for stochastic optimization, 3rd International Conference on Learning Representations, ICLR 2015 - Conference Track Proceedings (2015) 1–15.

37. F. Isensee, P. F. Jaeger, S. A. Kohl, J. Petersen, K. H. Maier-Hein, nnU-Net: a self-configuring method for deep learning-based biomedical image segmentation, Nature Methods 18 (2) (2021) 203–211. doi:10.1038/s41592-020-01008-z. URL http://dx.doi.org/10.1038/s41592-020-01008-z

38. A. Kazerouni, E. K. Aghdam, M. Heidari, R. Azad, M. Fayyaz, I. Hacihaliloglu, D. Merhof, Available online 23, Medical Image Analysis 88 (2023) 1361–8415. doi:10.1016/j.media.2023.102846. URL https://doi.org/10.1016/j.media.2023.102846

39. T. Park, A. A. Efros, R. Zhang, J.-Y. Zhu, Contrastive learning for unpaired image-to-image translation, in: Computer Vision–ECCV 2020: 16th European Conference, Glasgow, UK, August 23–28, 2020, Proceedings, Part IX 16, Springer, 2020, pp. 319–345.

40. S. Bohlender, I. Oksuz, A. Mukhopadhyay, A Survey on Shape-Constraint Deep Learning for Medical Image Segmentation, IEEE REVIEWS IN BIOMEDICAL ENGINEERING 16 (2021) 2023. doi:10.1109/RBME.2021.3136343. URL https://www.ieee.org/publications/rights/index.html

41. J. Ma, Y. He, F. Li, L. Han, C. You, B. Wang, Segment Anything in Medical Images, Tech. rep.

